# Barriers and enablers to obesity prevention in female-only high schools in Riyadh: A qualitative study exploring healthy eating, physical activity and school-based interventions using the COM-B Model

**DOI:** 10.1101/2025.11.05.25339463

**Authors:** Sarah Aldukair, Jayne V. Woodside, Khalid Almutairi, Laura McGowan

## Abstract

**Background:** In the Kingdom of Saudi Arabia (KSA), adolescent health is suboptimal. Findings reported 79% of youths aged 15–29 were physically inactive with 30% living with overweight or obesity. Poor dietary habits further complicate the obesity epidemic. Schools are promoted as key settings for obesity prevention, yet little is known about female-only high schools. This study explored barriers and enablers to healthy eating (HE), physical activity (PA), and obesity prevention school-based interventions (SBIs) through conducting focus group discussions (FGDs) with students and staff.

**Methods:** Nine FGDs were conducted across three female public high schools in Riyadh from varying deprivation levels; six with 37 students (aged 16–17) and three with 19 staff members. A semi-structured topic guide, informed by the COM-B model, explored capabilities, opportunities, and motivations related to obesity prevention. Framework analysis identified key barriers and enablers to HE, PA, and SBIs implementation.

**Results:** Barriers emerged across all COM-B constructs. Capability-related barriers included lack of trained staff. Opportunity-related barriers were most prominent, including hot weather, curriculum limitations, and built school environment. Staff and students collectively agreed that low student motivation was a key barrier. School staff highlighted structural enablers such as the physical education curriculum, while students identified individual-level motivators including willpower, improved mood, health, and body image. No mutual enablers were identified across staff and student groups.

**Conclusion:** Female-only high schools in KSA face major barriers to obesity prevention SBIs. Addressing these barriers through context-specific, multi-level approaches integrating staff and student perspectives is critical for effective SBIs.

**KEY MESSAGES:** *What is already known on this topic:*

- Childhood obesity rates in KSA are among the highest globally and driven by poor dietary habits, physical inactivity, socio-cultural and environmental factors.
- Schools are considered a key setting for health promotion.
- Obesity prevention initiatives within the context of female-only schools in KSA remains underexplored with most evidence originating from Western or mixed-gender settings.
- Female-only schools in KSA operate within unique cultural and environmental conditions (such as gender segregation and limited physical activity opportunities for females).
- This creates a need to further explore how to effectively design and implement SBIs in this unique context.

*What this study adds:*

- This is the first qualitative study to investigate barriers and enablers to healthy eating, physical activity and school-based obesity prevention interventions implementation among both female high school students and staff in Riyadh.
- By employing the COM-B model, the socio-ecological model and using framework analysis, the study offers a structured and behaviourally informed perspective.
- This study identifies structural and motivational barriers, such as the built school environment, lack of trained staff, curriculum limitations, and lack of student motivation for healthy eating and physical activity.
- It also identifies a range of behaviourally-informed enablers across COM (capability, opportunity and motivation) such as staff cooperation, peer support, the introduction of a physical education curriculum, body image, and health-related motivators.
- These findings provide context-specific understanding of obesity prevention SBIs in Saudi schools.

*How this study might affect research, practice, or policy:*

- This study highlights the need for culturally appropriate and context-specific obesity prevention SBIs, that address the individual, interpersonal, organizational, community, and public policy levels.
- It supports staff training, improvement of the built school environment, and practical PA in schools.
- Informs future SBIs research in KSA and gulf cooperation countries with similar contexts.

## INTRODUCTION

Childhood obesity is a major public health challenge globally and particularly in SA, reporting some of the highest rates of overweight and obesity (1). The 2019 KSA World Health Survey indicates that adolescent health is suboptimal, reporting 30% of 15–29-year-olds are living with overweight or obesity, with pre-diabetes and raised cholesterol affecting 10% and 39% of respondents, respectively alongside 79% having insufficient physical activity (PA) (2). Systematic reviews reveal that sedentary lifestyles and poor dietary behaviours are widespread among adolescents, including high fast-food consumption, low fruit and vegetable intake, breakfast skipping (3-5) and a 2023 scoping review illustrated that approximately 80–90% of adolescents fail to meet recommended daily PA targets (6).

The World Health Organization (WHO) identifies schools as key settings for promoting healthy behaviours (7-8). A qualitative systematic review of 18 school-based studies concluded that schools play a crucial role in obesity prevention by providing opportunities to improve PA and healthy eating (HE) behaviours, however, interventions are often limited in reach or efficacy (9). In KSA, little is known about the barriers and enablers to effective implementation of obesity prevention SBIs in female schools, where gender-segregation, cultural restrictions, and limited physical education (PE) provision until 2017 represent unique challenges (10-12). Female adolescents in KSA face additional barriers, including biological predispositions to adiposity (13) and social pressures related to body image (14-15). Although KSA’s Vision 2030’s recent policies aim to improve PA opportunities for females, cultural and environmental constraints remain (16).

Limited evidence about barriers and enablers to SBIs, HE, and PA from KSA exists, with most research being conducted in Western contexts (17-20). Some studies offer insights from Jeddah (21–23), but little is known about female high schools in Riyadh, particularly from students’ and school staff perspectives. Behaviour change models can offer valuable frameworks to conceptualise barriers and enablers to behaviour change, such as the COM-B model, which conceptualizes behaviour as the interaction of capability (physical skills and psychological knowledge), opportunity (physical resources and social support), and motivation (automatic habits and reflective intentions) (24). It can be used to guide context-specific intervention design, for example, the Healthy Buddies program employed this model to improve healthy behaviours in elementary schools (25). Additionally, given the complex myriad of factors which contribute to the development of obesity, the Social Ecological Model (SEM) was employed to provide a comprehensive perspective on obesity risk factors by recognizing the multiple layers of influence; individual, interpersonal, organizational, community, and public policy (26).This study explores barriers and enablers to HE and PA from students, alongside exploring barriers and enablers to implement obesity prevention SBIs from staff, guided by COM-B and the SEM.

## METHODS

### 1. Design of qualitative FGDs

This qualitative study was comprised of semi-structured FGDs with separate groups for school staff (school staff and principals) and high school students to explore barriers and enablers of HE, PA, and implementing obesity prevention SBIs across high schools situated in areas of varying economic deprivation. Students were drawn from grades 10-11, typically aged 16–17 years.

A semi-structured topic guide was developed informed by the COM-B model (24), exploring capabilities, opportunities, and motivations of school staff regarding barriers and enablers of implementing obesity prevention SBIs; and for students regarding barriers and enablers to HE and PA. FGDs were conducted separately for staff and students; topic guides are provided in supplementary material (i).

### 2. Personal and public involvement and engagement (PPIE)

Ensuring research is relevant and framed sensitively is important, especially when working with adolescents on the topic of HE and PA. Given the involvement of high school students in the present study, PPIE was incorporated to obtain adolescent input on the research design, framing and to assess their understanding of the FGD topic guide (27). Using the researcher’s local networks, a snowballing approach was taken to engage PPIE representatives. Two young people from the target age group (females) joined an informal video call with the researcher [SA] to provide feedback on the topic guide and framing of the research, and revisions were made accordingly. This ensured the study addressed student priorities and aligned with their language/terminology.

### 3. Setting, population, and sampling technique

There are 279 female governmental high schools in Riyadh. Schools were categorised into three deprivation areas; low deprivation schools (LDS), medium deprivation schools (MDS), and high deprivation schools (HDS). Deprivation status was determined based on the price per square metre of land, according to documents from the Saudi Real Estate General Authority (28).

Schools within each category were randomized in an Excel sheet and numbered, and three schools were drawn randomly from each list using random number generators. The first schools contacted from the low and medium deprivation status cohorts agreed to participate. After the first HDS declined participation in the study, a second school was randomly selected from the remaining list and agreed to participate. The FGDs were then conducted in these three female public high schools in Riyadh with differing levels of economic deprivation. Six separate FGDs were conducted with students (two per school) and three with staff (one per school). The target sample size for each FGD was 6-8 participants. Given the age of participants, the nature of the topic and the complexity of the issue being discussed, a smaller group was considered more desirable (29). Eligible participants included female students aged 16-17 years and school staff at the selected schools who were available during the scheduled FGDs.

### 4. FGDs participants and recruitment

Eligible students were provided with a participant information sheet (PIS) and parental consent forms; participants were randomly selected alphabetically by surname. Staff received a PIS and were invited to participate based on availability and consent.

Each FGD lasted approximately one hour, conducted in Arabic by the lead researcher [SA]. Recordings were transcribed verbatim, translated into English, and back-translated for accuracy by [SA] and verified by a certified translator.

### 5. Data collection and storage

FGDs were audio-recorded anonymously using a password-protected device, with recordings saved in encrypted folders labeled by school, deprivation level, and participant type. The master list was kept separately to ensure anonymity.

### 6. Qualitative analysis

The lead researcher [SA] initially read the transcripts repeatedly, and generated codes. Double-coding and discussions with LM and the supervisory team ensured triangulation and consensus on the coding approach. Data were analyzed according to COM-B constructs: physical and psychological capability, physical and social opportunity, and automatic and reflective motivation. Codes were categorised as barriers or enablers. A framework analysis was employed to guide the qualitative analysis of FGD (30). The primary aim of framework analysis is to recognize, describe, and identify prominent trends present in the qualitative data under investigation and organize them according to the selected model, COM-B, supported by NVivo 12 software.

### 7. Reflexivity

The research team’s expertise shaped study design and interpretation. Dr. Laura McGowan has training in psychology, health psychology, obesity and health behaviour change; Prof. Jayne Woodside in nutrition and public health; and Sarah Aldukair in public health and health education.

### 8. Ethical considerations

The study was approved by Princess Nourah bint Abdulrahman University institutional review board and Queen’s University Belfast’s Faculty of Medicine, Health and Life Sciences REC (MHLS 23_62). The Saudi Ministry of Education granted permission to access schools. The ethical approval conformed to the principles embodied in the Declaration of Helsinki. Due to potential sensitivity of HE, PA, and weight-related discussions, participants were provided with information about support services. Participation was voluntary, with written informed consent obtained from staff, students, and parents prior to participation.

## RESULTS

Three high schools were recruited to the FGD study and are outlined in TABLE.1. A total of 37 female students and 19 staff members (including teachers and principals) participated across the nine FGDs. Findings are presented using the COM-B framework, highlighting barriers and enablers to HE, PA, and the implementation of obesity prevention SBIs as highlighted in TABLE.2.

**TABLE.1:** Number of FGD participants recruited into the study.

| **School area** | **Number of FGD with students** | **Number of FGD with school staff** | **Number of FGD participants achieved** |
| --- | --- | --- | --- |
| 1 LDS* | 2 | 1 | Students (n= 13 across two FGDs)<br>School staff (n= 6) |
| 1 MDS† | 2 | 1 | Students (n= 12 across two FGDs)<br>School staff (n= 7) |
| 1 HDS‡ | 2 | 1 | Students (n= 12 across two FGDs)<br>School staff (n= 6) |
| **Total across 3 different schools from 3 different areas of deprivation** | **9 FGD** |  | **37 students, 19 school staff members** |

\* Low deprivation school (LDS)
† Medium deprivation school (MDS)
‡ High deprivation school (HDS)

**TABLE.2:**
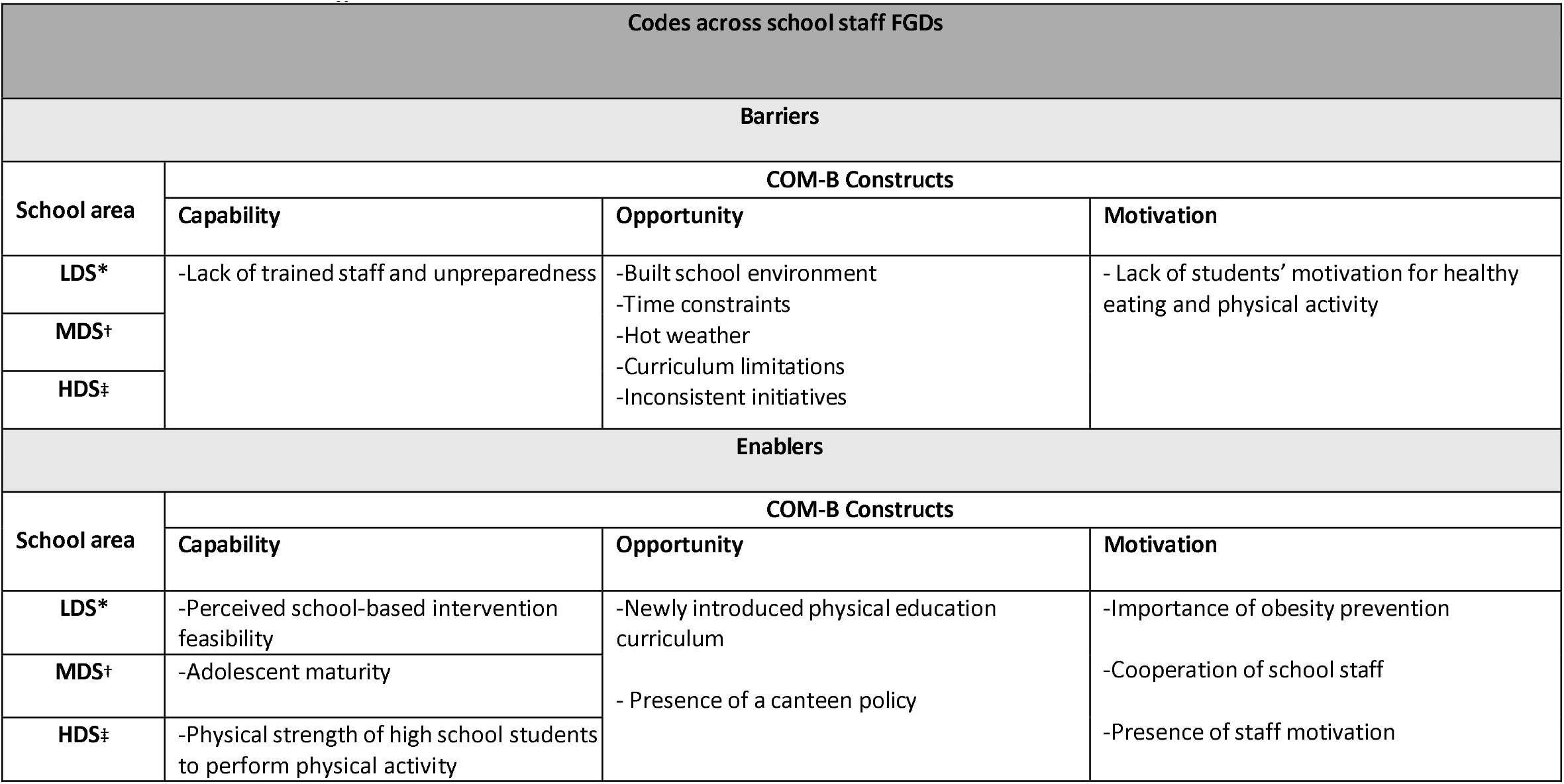
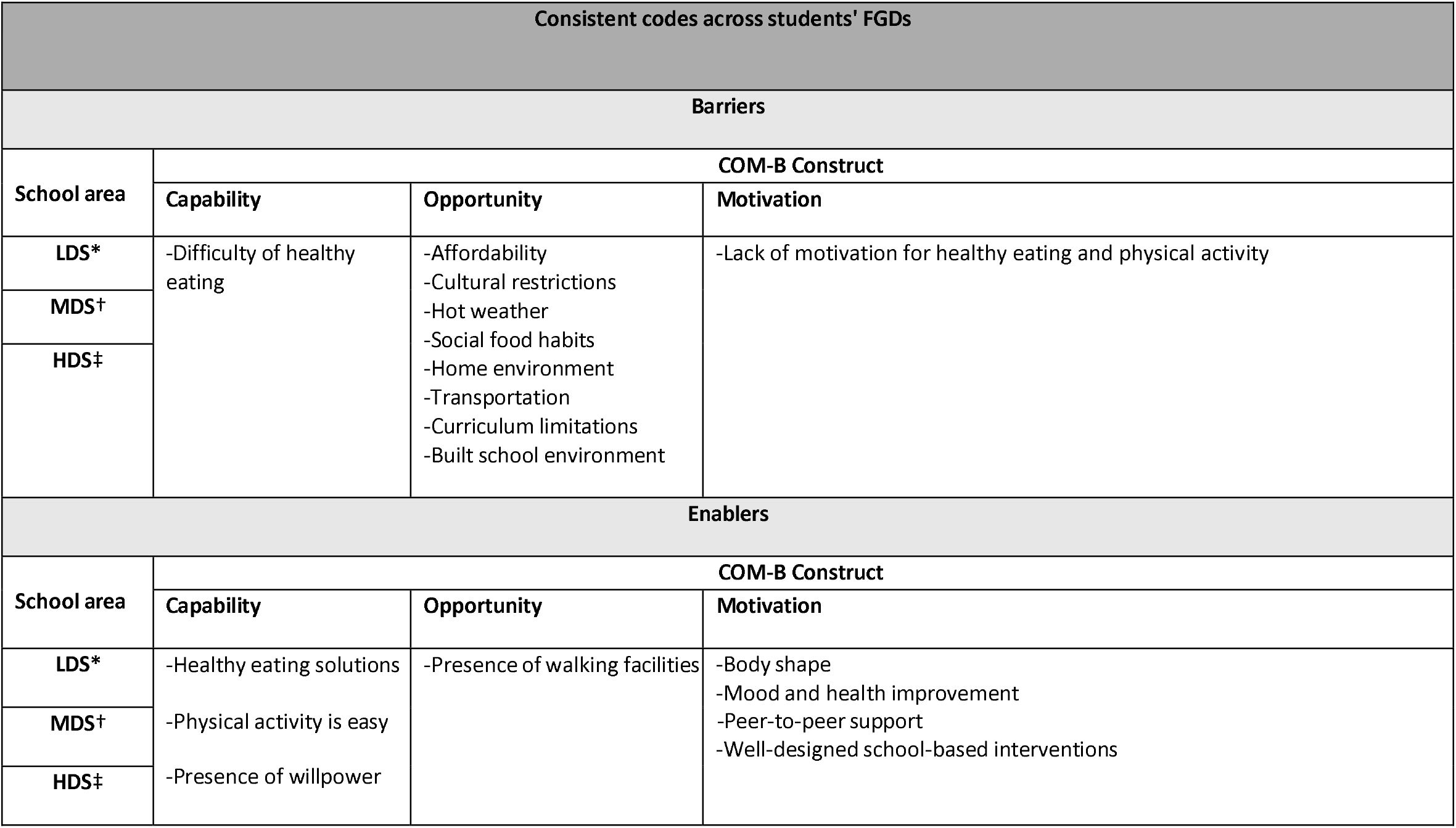
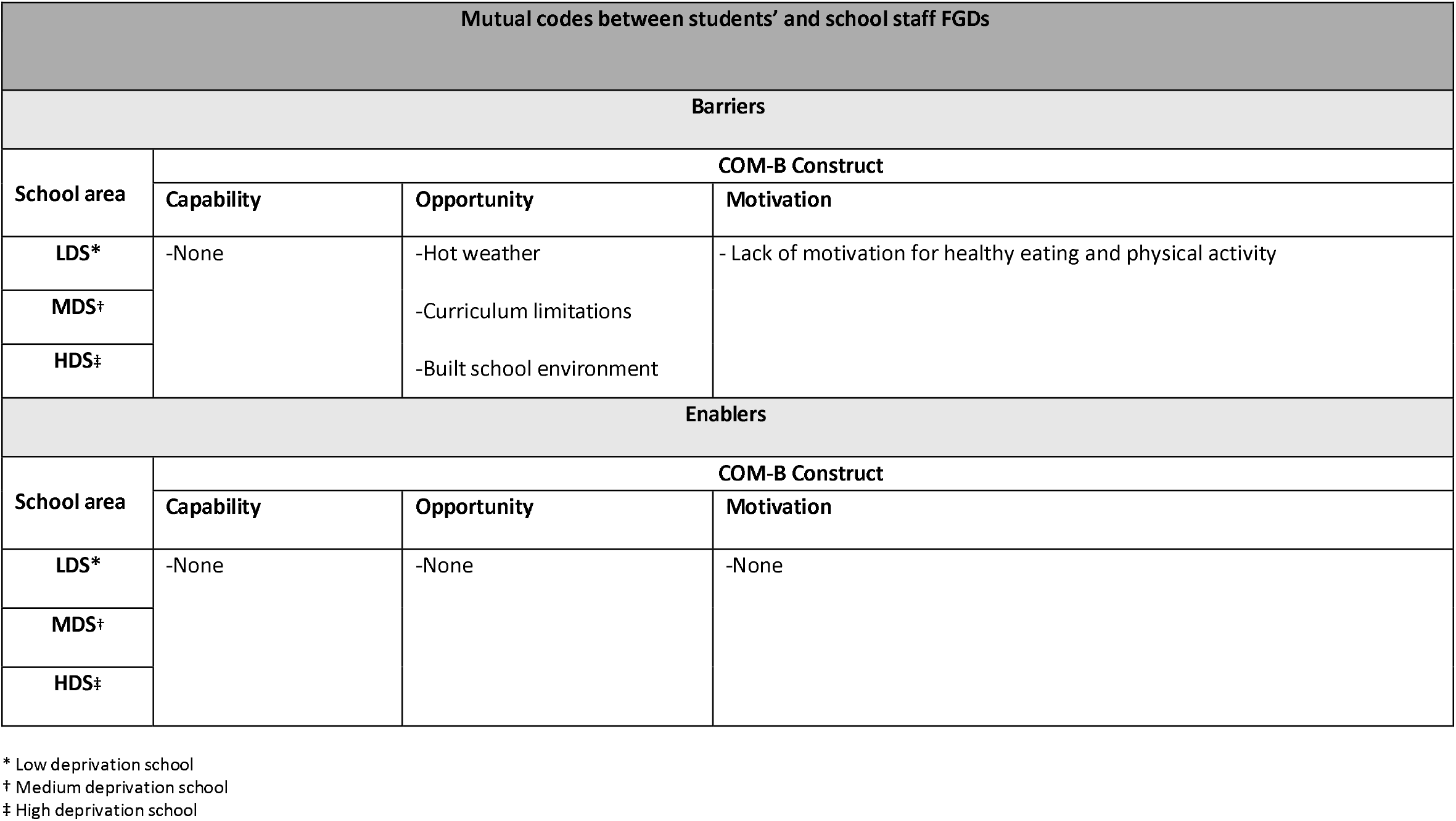
Codes across school staff and students’ FGDs.

### 1. School staff perspectives

#### 1.1 Barriers and enablers relating to *capability*

In the school staff FGDs, factors relating to capability were consistent across the different schools sampled, with lack of trained staff noted as a prominent barrier.

> *“I have no experience in the field of school-based interventions*… *I am supposed to be familiar with the subject by taking a short course or training before the beginning of the intervention*.*”*
>
> *(Teacher 2-HDS)*

There was less consistency regarding enablers, although each school identified a different enabler; perceived SBIs feasibility in the LDS, adolescent maturity in the MDS, and physical strength in the HDS.

> *“It is simple; provide fruits, vegetables, and healthy options in the canteen. I don’t think it will be difficult at all*…*” (Teacher 1-LDS)*

> *“High school students are older than middle or primary school students, so they would be more convinced in obesity interventions because they are always concerned with their body shape, and this can motivate them to be healthier because they want to look beautiful*.*” (Teacher 2-MDS)*

> *“*…*students at this age are young and have the physical strength to be active and practice physical activity, but they need a suitable environment for that*.*” (Principal-HDS)*

#### 1.2 Barriers and enablers relating to *opportunity*

Factors relating to opportunity were consistent across different schools; with the built school environment as a prominent barrier. The majority of school staff discussed the barriers to PA and HE related to the opportunity of physical environment:

> *“The school environment is not ready at all, at all, at all! Not even the slightest readiness*… *How can we implement a school program in our school? The environment is not supportive of health programs*… *because canteens are very unhealthy and there is no gym*… *the situation of our canteen can hinder positive change, no matter how good the intervention is*.*’’ (Teacher 6-MDS)*

> *“Let’s say the school makes efforts to support healthy choices, but when the student looks at the school environment that contradicts what has been taught, school efforts are ruined. The students may be aware of the importance of healthy eating and physical activity, but the school environment does not support their ideas*.*” (Teacher 4-HDS)*

The newly introduced physical education (PE) curriculum emerged as a consistent enabler across different schools:

> *“The newly introduced physical education curriculum is an excellent addition for the students”-*
>
> *(Teacher 1-LDS)*

> *“The PE curriculum is a nice addition*… *and hopefully soon it will include exercising sessions*.*”*
>
> *(Teacher 3-MDS)*

#### 1.3 Barriers and enablers relating to *motivation*

Factors relating to motivation were also consistent across schools, with the importance of obesity prevention identified as an enabler:

> *“I believe that healthy eating and physical activity are very important, especially among teenagers, so believing in the importance of obesity prevention motivates me*…*” (Teacher 1-HDS)*

> *“The importance of obesity prevention, healthy eating and physical activity, especially for adolescent girls*… *I strongly believe that maintaining a healthy lifestyle is one of the most important things*.*” (Principal-LDS)*

Lack of student motivation, however, was highlighted as a barrier:

> *“I would like to add that it is something that entirely depends on the student’s beliefs and motivation*…*students are not motivated to eat healthy or exercise” (Teacher 3-HDS)*

> *“I try to motivate my students, but the students are not cooperative with me, and I think the biggest reason is being shy to perform physical activity*.*” (Teacher 4-LDS)*

### 2. Students’ perspectives

#### 2.1 Barriers and enablers relating to *capability*

In the student FGDs, factors relating to capability were consistent across schools, with difficulty of HE due to taste preferences as the main barrier:

> *“It is very difficult to eat healthy because I have to cut the sugars and refrain from foods I like. Healthy food is not tasty so it will be hard for me to eat it*.*” (Student 1-MDS)*

> *“For me following a healthy diet is extremely difficult because I have noticed that my mood is connected to food immensely*… *For example, if there is salad and burger what would you choose? For sure all of us will choose the burger, it tastes better!” (Student 4-LDS)*

Enablers were consistent across schools and included awareness of HE solutions, recognition that PA can be simple, and the presence of ‘willpower”” or motivation was considered important from their perspective:

> *“If eating healthy it is a gradual diet and there is no sudden abstinence from unhealthy foods, it will become less difficult*.*” (Student 1-MDS)*

> *“I don’t consider physical activity difficult; it can be for 15 minutes a day only*… *the most important thing is to devote time to it*.*” (Student 2-HDS)*

> *“If a person has the determination and willpower, physical activity will become easy*… *and gradually it will become doable” (Student 1-LDS)*

#### 2.2 Barriers and enablers relating to opportunity

Factors relating to opportunity were also consistent across schools; barriers included affordability, cultural restrictions, hot weather, social food habits, unsupportive home environments, transportation challenges, curriculum limitations, and inadequate built school environments:

> *“Healthy food is expensive and unhealthy food is available and cheap*… *Healthy alternatives are always expensive*.*” (Student 5-LDS)*

> *“It is socially unacceptable for a girl to go out to walk in the neighborhood” (Student 5-HDS)*

> *“I like walking and I don’t think it is hard but it needs a nice weather and the weather in our country is hot almost all year long*.*” (Student 6-MDS)*

> *“In social gatherings it is hard to refuse the food served because it is considered impolite*… *There is a lot of pressure to eat at social events and the food is extremely unhealthy” (Student 4-LDS)*

> *“At home, vegetables and fruits aren’t always available. Healthy food isn’t available*.*” (Student 2-LDS)*

> *“Walkways are nearby, but we can’t just walk, we must drive to that place*… *which is an obstacle, transportation is not always available*.*” (Student 3-LDS)*

> *“Although we study physical education but the problem is we don’t apply it. We only study it theoretically*.*” (Student 1-MDS)*

> *“The canteen is extremely unhealthy, and we don’t do any physical activity*.*” (Student 1-LDS)*

The presence of walking facilities was identified as a consistent enabler across different schools:

> *“I like walking and I don’t think it is hard, but it needs a nice weather and the weather in our country is hot almost all year long. However, there are alternatives like the air-conditioned malls*.*” (Student 3-MDS)*

#### 2.3 Barriers and enablers relating to *motivation*

Factors relating to motivation were consistent across schools, with the lack of motivation to engage in HE and PA as a barrier.

> *“I just don’t have the motivation to prevent myself from eating unhealthy food*.*” (Student 1-HDS)*

> *“I have no motivation to eat healthy or exercise” (Student 3-LDS)*

Consistent enablers included the importance of body shape, mood and health improvement, peer-to-peer support, and well-designed SBIs that include competitions.

> *“When my body shape becomes nice, I feel that I have succeeded*.*” (Student 6-MDS)*

> *“When I eat healthy food and exercise that would elevate my physical and mental health. Also, when that reflects on my skin, my hair and my looks*.*” (Student 1-HDS)*

> *“If we all eat healthy food together as friends, we will certainly get excited*.*” (Student 1-LDS)*

> *“There should also be competitions between schools to boost competitiveness between students” (Student 6-MDS)*

### 3. Mutual findings

Across both groups, mutual barriers were consistently identified; extreme heat, curriculum limitations, built school environment, and the lack of students’ motivation for HE and PA. No mutual enablers were identified across both groups.

### 4. Summary of barriers and enablers mapped to the SEM

COM-related barriers and enablers emerged at multiple levels of the SEM: *individual* (lack of motivation, difficulty of HE, hot weather, lack of trained staff, presence of willpower, physical strength, HE solutions, body shape, adolescent maturity, PA is easy and mood and health improvement), *interpersonal* (lack of staff motivation, social food habits, cooperation of school staff, peer support and SBIs are simple to implement), *institutional* (curriculum limitations, inconsistent initiatives, built school environment, newly introduced PE curriculum, presence of canteen policy and well-designed SBIs), *community* (social food habits, transportation, home environment, cultural restrictions, walking facilities and importance of obesity prevention), and *policy* (affordability). This multi-level perspective highlights the importance of addressing obesity influences beyond the individual-level to achieve behavioural change, as illustrated in *FIGURE.1*:

**FIGURE.1:**
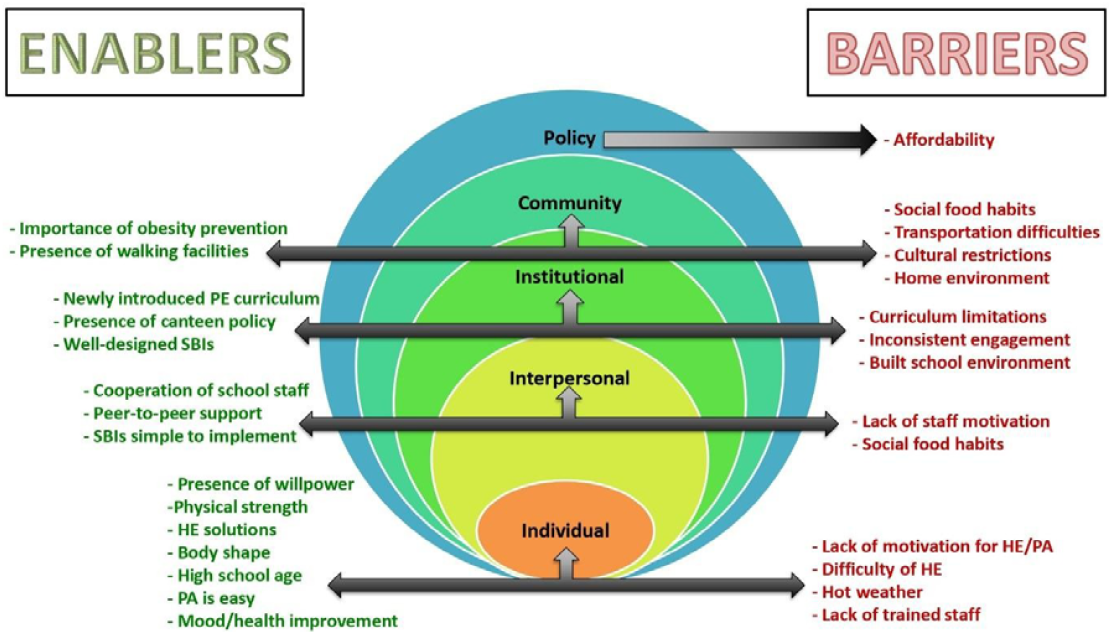
Summary of barriers and enablers influencing HE, PA and SBIs implementation mapped across the SEM (26)

## DISCUSSION

This study identified key capability, opportunity and motivation-related barriers and enablers to implementing obesity prevention SBIs targeting HE and PA among female students and staff in Riyadh, KSA. In school staff FGDs, key barriers included lack of trained staff (capability), curriculum limitations, built school environment, time constraints, hot weather, inconsistent initiatives (opportunity), and low student motivation for HE and PA (motivation). These barriers were consistent across schools of differing deprivation levels. Enablers included student age and strength, perceived SBIs feasibility, newly introduced physical education curriculum, canteen policy, staff cooperation and motivation.

In student FGDs, barriers to HE and PA included difficulty and affordability of HE, cultural and environmental factors, transportation, and lack of motivation. Enablers included practical HE solutions, ease of PA, presence of willpower, presence of walking facilities, body image, mood and health improvement, peer support, and well-designed SBIs. Both barriers and enablers were consistent across schools from varying deprivation levels. Both groups identified mutual barriers: hot weather, curriculum limitations, poor school infrastructure, and low student motivation. No mutual enablers were found.

The key findings align with previous research emphasizing the environmental, structural and motivational barriers to HE and PA in schools (17-20), while also emphasizing context-specific issues relating to Saudi female schools, such as gender-segregated environments, unsupportive school infrastructures, the availability of PA, and policies unique to female students (15). This study adds to the literature by highlighting the perspectives of both students and staff, reinforcing the need for tailored, multi-level interventions.

Despite evidence on the effectiveness of SBIs to prevent obesity (7-9), a significant number of schools fail to successfully implement effective interventions due to several barriers (31). This study highlights barriers and enablers in Saudi female high schools, underscoring the need for context-specific approaches to strengthen schools’ role in obesity prevention, which go beyond targeting education alone and focus on multiple capability, opportunity and motivational strategies to improve HE and PA behaviours.

School staff identified the lack of training, heavy workloads, and time constraints as barriers to the implementation of obesity prevention SBIs. This aligns with previous findings identifying time constraints as a major barrier to implementing effective SBIs (32). School staff and students jointly agreed that the newly introduced physical education curriculum was frustrating as it only contained theoretical information, stressing the need for an applied component to make it more useful. This is consistent with findings from a systematic review revealing that implementation of diverse PA activities enhances intervention impact (33). The built school environment was criticized for lacking adequate PA facilities and HE offerings, which aligns with two Saudi studies revealing organizational challenges in creating school environments conducive to PA and HE (21-22). Extreme weather was also a major barrier to PA, given the open structure of Saudi school buildings and the dangers of exercising outdoors in the heat.

Indeed, previous research conducted in Brazil has shown that poor weather was one of the main perceived barriers to PA among adolescents (34). Finally, both school staff and students highlighted low student motivation as a persistent barrier, compounded by the lack of sustained health initiatives, often limited to one-time events.

Despite the presence of several barriers, school staff identified enablers. Enablers varied by deprivation level; In the LDS, staff perceived SBIs as feasible to implement, though systemic barriers remained. In the MDS, adolescent maturity was seen as an enabler, since students are more health and appearance conscious. In the HDS, staff highlighted students’ physical strength, provided the environment supports PA. Across all schools, staff valued the introduction of the PE curriculum, the canteen policy, the importance of obesity prevention, and their own cooperation and motivation as enablers to support future interventions.

Students consistently identified the difficulty of HE as a major barrier, linking it to taste preferences, affordability, and unsupportive home environments. Cultural restrictions were reported as barriers to PA, particularly the limitation on females walking alone in neighborhoods. Previous research explicitly identified cultural restrictions as barriers to PA among Saudi females (12). Students also identified social gatherings as barriers to healthy eating, consistent with previous research revealing that such settings often hinder HE among young people (35). Transportation challenges limited access to PA facilities, reflecting Riyadh’s car-dependent infrastructure. Extreme heat further discouraged outdoor PA. These barriers highlight the influence of cultural, environmental, and structural factors on adolescent health behaviours.

Conversely, students identified several enablers to healthier behaviours. They highlighted dietary solutions such as moderation, gradual diet changes, and provision of healthy options in schools as enablers to HE. This supports earlier findings that offering healthy food in school canteens enables HE (21). While HE was perceived as difficult, PA was seen as relatively easy if time, determination and willpower were present. The presence of walking facilities, such as in neighborhood walkways and in malls, were considered supportive environments enabling PA. Students also highlighted body image, mood, and health improvement as key motivators to HE and PA, aligning with previous findings that young adults are motivated by appearance, wellbeing, and health-related factors when making dietary choices (36). Similarly, findings from a qualitative study revealed that motivators for PA included both body image and health improvement among Saudi females (12). Peer support was emphasized as a powerful enabler, with students noting that engaging in HE and PA with friends was more motivating than doing so alone. Indeed, peer support has shown positive effects on HE and PA behaviours (37). Finally, students criticized existing health programs as overly theoretical and called for well-designed SBIs that are practical, engaging, and skill focused.

Although barriers to HE and PA are often more significant in high deprivation settings, this study and prior research (38) demonstrate that such challenges are present even in LDS, where unhealthy canteen options, limited availability of healthy foods, and affordability issues represent structural issues affecting different socioeconomic groups.

A key strength of this study is its novelty as the first qualitative study to investigate barriers and enablers to HE, PA, and SBIs implementation among female high school students and staff in Riyadh, guided by behavior change theories. The inclusion of schools from different deprivation levels captured diverse perspectives. Guided by the COM-B model, the SEM and using framework analysis, the study offers a structured and behaviourally informed perspective on future targets for SBIs in this setting and context, beyond the level of the individual. The qualitative approach captured lived experiences and insights not accessible through quantitative methods.

Limitations include the small sample size and number of schools included, which limits generalisability to other populations such as male students, private schools, or schools in different regions. Despite this, findings provide valuable insights for schools in KSA and the wider Gulf region with similar socio-cultural contexts.

Future research should consider the inclusion of male students, other school staff such as canteen providers and MOE stakeholders, and making a comparison between private and public schools across different regions. Further investigation of cultural barriers to PA among Saudi adolescents is also recommended. These future directions will support the development of tailored, multi-level obesity prevention SBIs in Saudi schools.

## CONCLUSION

School staff and students in female-only KSA high schools across the socio-economic spectrum reported numerous barriers and enablers across COM-B constructs, highlighting the need for comprehensive obesity prevention SBIs to address the lack of PA and HE provisions in female public schools. However, female high schools face significant challenges regarding a school environment that supports the implementation of obesity prevention SBIs that promote HE and PA. In order to implement obesity prevention SBIs aimed at improving HE and PA behaviours in high schools, policy changes should address organizational challenges that Saudi schools are faced with, support school staff with adequate training, consider the socio-cultural context of KSA, and advocate for policies to improve the school environment and curriculum. Understanding the school context will help support the development of future obesity prevention SBIs.

## DECLARATIONS

### Ethical approval

Approved by Princess Nourah bint Abdulrahman University’s institutional review board, Saudi Ministry of Education, and Queen’s University Belfast REC (MHLS 23_62). Written consent was obtained from all participants.

### Data availability

All data produced in the present study are available upon reasonable request to the authors.

### Competing interests

The authors declare no competing interests

### Funding

This study was funded by Princess Nourah bint Abdulrahman University as part of a doctoral PhD undertaken at Queen’s University Belfast.

### Authors’ contributions

SA, LM and JW designed the study, SA conducted the study, collected and analysed data, and drafted the initial manuscript. LM contributed to coding, analysis, and interpretation. JW and KA supervised study design and JW and LM provided critical revisions. All authors approved the final manuscript.

## Acknowledgements

We thank the participating schools, students, and staff for their valuable contributions, especially the PPIE contributors.

## Notes

### Competing Interest Statement

The authors have declared no competing interest.

### Funding Statement

This study was funded by Princess Nourah bint Abdulrahman University as part of a doctoral PhD undertaken at Queens University Belfast.

### Author Declarations

Ethics committee/institutional review board of Princess Nourah bint Abdulrahman University and Saudi Ministry of Education and Queens University Belfast REC (MHLS 23_62) gave ethical approval for this work. Written consent was obtained from all participants.

## REFERENCES

(1) NCD Risk Factor Collaboration. (2017). Worldwide trends in body-mass index, underweight, overweight, and obesity from 1975 to 2016: A pooled analysis of 2416 population-based measurement studies in 128·9 million children, adolescents, and adults. The Lancet, 390(10113), 2627–2642. 10.1016/S0140-6736(17)32129-3

(2) World Health Survey Saudi Arabia KSAWHS, 2019 [Internet]. Riyadh; 2019 [cited 3 September 2023]. Available from: https://www.moh.gov.sa/en/Ministry/Statistics/Population-Health-Indicators/Documents/World-Health-Survey-Saudi-Arabia.pdf

(3) Al-Hazzaa HM. Physical inactivity in Saudi Arabia revisited: A systematic review of inactivity prevalence and perceived barriers to active living. International journal of health sciences. 2018 Nov;12(6):50.

(4) Mumena WA, Ateek AA, Alamri RK, Alobaid SA, Alshallali SH, Afifi SY, Aljohani GA, Kutbi HA. Fast-food consumption, dietary quality, and dietary intake of adolescents in Saudi Arabia. International journal of environmental research and public health. 2022 Nov 16;19(22):15083.

(5) Alasqah I, Mahmud I, East L, Usher K. Patterns of physical activity and dietary habits among adolescents in Saudi Arabia: A systematic review. Int J Health Sci (Qassim). 2021 Mar-Apr;15(2):39-48. PMID: 33708043; PMCID: PMC7934132.

(6) Evenson KR, Alhusseini N, Moore CC, Hamza MM, Al-Qunaibet A, Rakic S, Alsukait RF, Herbst CH, AlAhmed R, Al-Hazzaa HM, Alqahtani SA. Scoping Review of Population-Based Physical Activity and Sedentary Behaviour in Saudi Arabia. J Phys Act Health. 2023 Apr 25;20(6):471–486. doi: 10.1123/jpah.2022-0537. PMID: 37185448.

(7) World Health Organization, UNESCO. Making every school a health-promoting school: Global standards and indicators [Internet]. Geneva: World Health Organization; 2021 [cited 2025 Oct 11]. Available from: https://cdn.who.int/media/docs/default-source/health-promotion/9789240025059-eng.pdf

(8) Singhal J, Herd C, Adab P, Pallan M. Effectiveness of school-based interventions to prevent obesity among children aged 4 to 12 years old in middle-income countries: a systematic review and meta-analysis. Obesity reviews. 2021 Jan;22(1):e13105.

(9) Clarke J, Fletcher B, Lancashire E, Pallan M, Adab P. The views of stakeholders on the role of the primary school in preventing childhood obesity: a qualitative systematic review. Obesity Reviews. 2013 Dec;14(12):975–88.

(10) Aljaaly EA. Physical activities of young girls in Jeddah, Saudi Arabia. Arab Journal of Nutrition and Exercise (AJNE). 2016:122–30.

(11) Mobaraki AE, Soderfeldt B. Gender inequity in Saudi Arabia and its role in public health. EMHJ-Eastern Mediterranean Health Journal, 16 (1), 113–118, 2010. 2010.

(12) Aljehani N, Razee H, Ritchie J, Valenzuela T, Bunde-Birouste A, Alkhaldi G. Exploring female university Students’ participation in physical activity in Saudi Arabia: a mixed-methods study. Frontiers in public health. 2022 Mar 18;10:829296.

(13) Taylor RW, Williams SM, Carter PJ, Goulding A, Gerrard DF, Taylor BJ. Changes in fat mass and fat-free mass during the adiposity rebound: FLAME study. International Journal of Pediatric Obesity. 2011 Jun 1;6(Sup3):e243-251.

(14) Elia C, Karamanos A, Silva MJ, O’Connor M, Lu Y, Dregan A, Huang P, O’Keeffe M, Cruickshank JK, Enayat EZ, Cassidy A. Weight misperception and psychological symptoms from adolescence to young adulthood: longitudinal study of an ethnically diverse UK cohort. BMC Public Health. 2020 Dec;20(1):1–4.

(15) Glasofer DR, Haaga DA, Hannallah L, Field SE, Kozlosky M, Reynolds J, Yanovski JA, Tanofsky-Kraff M. Self-efficacy beliefs and eating behaviour in adolescent girls at-risk for excess weight gain and binge eating disorder. International Journal of Eating Disorders. 2013 Nov;46(7):663–8.

(16) Saudi Vision 2030. National Transformation Program 2030. (2018). Available online at: https://www.vision2030.gov.sa/ (accessed 9 May 2024).

(17) Nathan N, Elton B, Babic M, McCarthy N, Sutherland R, Presseau J, Seward K, Hodder R, Booth D, Yoong SL, Wolfenden L. Barriers and facilitators to the implementation of physical activity policies in schools: a systematic review. Preventive medicine. 2018 Feb 1;107:45–53.

(18) Hayes CB, O’shea MP, Foley-Nolan C, McCarthy M, Harrington JM. Barriers and facilitators to adoption, implementation and sustainment of obesity prevention interventions in schoolchildren–a DEDIPAC case study. BMC Public Health. 2019 Dec;19:1–3.

(19) Payán DD, Sloane DC, Illum J, Farris T, Lewis LB. Perceived barriers and facilitators to healthy eating and school lunch meals among adolescents: a qualitative study. American journal of health behaviour. 2017 Sep 1;41(5):661–9.

(20) Martins J, Marques A, Sarmento H, Carreiro da Costa F. Adolescents’ perspectives on the barriers and facilitators of physical activity: a systematic review of qualitative studies. Health education research. 2015 Oct 1;30(5):742–55.

(21) Almughamisi M, O’Keeffe M, Harding S. Adolescent obesity prevention in Saudi Arabia: co-identifying actionable priorities for interventions. Frontiers in Public Health. 2022 May 10;10:863765.

(22) Almutairi N, Burns S, Portsmouth L. Barriers and enablers to the implementation of school-based obesity prevention strategies in Jeddah, KSA. International Journal of Qualitative Studies on Health and Well-being. 2022 Dec 31;17(1):2135197.

(23) Almutairi N, Burns S, Portsmouth L. Physical activity knowledge, attitude, and behaviours among adolescents in the Kingdom of Saudi Arabia prior to and during COVID-19 restrictions. J Obes. 2022;2022:1892017. doi:10.1155/2022/1892017.

(24) West R, Michie S. A brief introduction to the COM-B Model of behaviour and the PRIME Theory of motivation [v1]. Qeios. 2020 Apr 7.

(25) Nickel NC, Doupe M, Enns J, Santos RG. Differential effects of a school-based obesity prevention program: A cluster randomized trial. Matern Child Nutr. 2020;16(1):e13009. Available from: 10.1111/mcn.13009

(26) Elder JP, Lytle L, Sallis JF, Young DR, Steckler A, Simons-Morton D, Stone E, Jobe JB, Stevens J, Lohman T, Webber L. A description of the social–ecological framework used in the trial of activity for adolescent girls (TAAG). Health education research. 2007 Apr 1;22(2):155–65.

(27) Pizzo E, Doyle C, Matthews R, Barlow J. Patient and public involvement: how much do we spend and what are the benefits?. Health expectations. 2015 Dec;18(6):1918–26.

(28) Annual Reports [Internet]. Available from: https://rega.gov.sa/en/the-library/annual-reports/

(29) Ritchie J, Lewis J, Nicholls CM, Ormston R. Qualitative research practice. London: sage; 2003.

(30) Furber C. Framework analysis: a method for analysing qualitative data. African Journal of Midwifery and Women’s health. 2010 Apr;4(2):97–100.

(31) Fathi B, Allahverdipour H, Shaghaghi A, Kousha A, Jannati A. Challenges in developing health promoting schools’ project: application of global traits in local realm. Health promotion perspectives. 2014;4(1):9.

(32) St Leger, L., 2001. Schools, health literacy and public health: possibilities and challenges. Health Promotion International, 16, pp.197–205.

(33) Yuksel, H.S., Şahin, F.N., Maksimovic, N., Drid, P. and Bianco, A., 2020. School-based intervention programs for preventing obesity and promoting physical activity and fitness: a systematic review. International Journal of Environmental Research and Public Health, 17(1), p.347.

(34) Dambros, D.D., Lopes, L.F. and Santos, D.L., 2011. Perceived barriers and physical activity in adolescent students from a Southern Brazilian city. Revista Brasileira de Cineantropometria & Desempenho Humano, 13, pp.422–428.

(35) Neely, E., Walton, M. and Stephens, C., 2014. Young people’s food practices and social relationships: a thematic synthesis. Appetite, 82, pp.50–60.

(36) Munt AE, Partridge SR, Allman-Farinelli M. The barriers and enablers of healthy eating among young adults: a missing piece of the obesity puzzle. Nutrients. 2017;9(9):873.

(37) Mellanby, A.R., Rees, J.B. and Tripp, J.H., 2000. Peer-led and adult-led school health education: a critical review of available comparative research. Health Education Research, 15(5), pp.533–545.

(38) Yoong SL, Nathan NK, Wyse RJ, Preece SJ, Williams CM, Sutherland RL, et al. Assessment of the school nutrition environment: a study in Australian primary school canteens. Am J Prev Med. 2015 Aug;49(2):215–22. doi:10.1016/j.amepre.2015.02.002. PMID: 26091931.

