## Supplementary material (i) for "Barriers and enablers to obesity prevention in female-only high schools in Riyadh: A qualitative study exploring healthy eating, physical activity and school-based interventions using the COM-B Model"

***TABLE.1: School staff FGDs topic guide informed by the COM-B model***

| Type of FGD | COM-B Constructs | | |
|---|---|---|---|
| | **Capability** | **Opportunity** | **Motivation** |
| **With school staff (teachers/principals) to explore barriers of and enablers to implementing obesity prevention school-based interventions** | 1- How would you rate the school's capability and preparedness to implement obesity interventions?<br><br>2- How would you rate your capability of implementing an obesity school-based intervention? | 1-What are the barriers of implementing school-based obesity interventions?<br><br>2-What are the facilitators of implementing school-based obesity interventions?<br><br>3-Can you describe whether the school environment supports making healthy lifestyle choices?<br><br>4-What are the opportunities given to you by the Ministry of Education to encourage students to adopt healthy lifestyle practices? | 1-What motivates you to accept the implementation of a school-based obesity intervention?<br><br>2-What motivates you to encourage students to adopt healthy lifestyles? |

*TABLE.2: Students' FGDs topic guide informed by the COM-B model*

| Type of FGD | COM-B Constructs | | |
|---|---|---|---|
| | **Capability** | **Opportunity** | **Motivation** |
| **With high school students to explore barriers and enablers to healthy eating and physical activity** | 1- How easy or difficult is it to perform physical activity and why?<br><br>2- How easy or difficult is it to eat healthy foods and why?<br><br>3- When it comes to being successful in engaging in healthy behaviours like eating healthy nutritious food and being physically active, how do you define success? | 1- What are the social barriers to maintaining a healthy diet?<br><br>2- What are the physical barriers to maintaining a healthy diet?<br><br>3- What are the social barriers to being physically active?<br><br>4- What are the physical barriers to being physically active?<br><br>5- Can you describe whether your school/home/wider environment supports making healthy lifestyle choices? | 1- What motivates you to eat healthy and be physically active?<br><br>2- Can you give us some advice on how to promote healthy dietary behaviours among students?<br><br>3- How can a school-based intervention motivate you to adopt a healthy lifestyle? |
